# The impact of temperature and absolute humidity on the coronavirus disease 2019 (COVID-19) outbreak - evidence from China

**DOI:** 10.1101/2020.03.22.20038919

**Authors:** Peng Shi, Yinqiao Dong, Huanchang Yan, Xiaoyang Li, Chenkai Zhao, Wei Liu, Miao He, Shixing Tang, Shuhua Xi

**Author notes:** Correspondence to: Prof Shuhua Xi, Department of Environmental Health, School of Public Health, China Medical University, No. 77 Puhe Road, Shenyang North New Area, Shenyang, Liaoning Province, China. Peng Shi, Yinqiao Dong, Huanchang Yan, Xiaoyang Li, and Chenkai Zhao contributed equally to this work.

## Abstract

**OBJECTIVE:** To investigate the impact of temperature and absolute humidity on the coronavirus disease 2019 (COVID-19) outbreak.

**DESIGN:** Ecological study.

**SETTING:** 31 provincial-level regions in mainland China.

**MAIN OUTCOME MEASURES:** Data on COVID-19 incidence and climate between Jan 20 and Feb 29, 2020.

**RESULTS:** The number of new confirm COVID-19 cases in mainland China peaked on Feb 1, 2020. COVID-19 daily incidence were lowest at -10 °C and highest at 10 °C, while the maximum incidence was observed at the absolute humidity of approximately 7 g/m^3^. COVID-19 incidence changed with temperature as daily incidence decreased when the temperature rose. No significant association between COVID-19 incidence and absolute humidity was observed in distributed lag nonlinear models. Additionally, A modified susceptible-exposed-infectious-recovered (M-SEIR) model confirmed that transmission rate decreased with the increase of temperature, leading to further decrease of infection rate and outbreak scale.

**CONCLUSION:** Temperature is an environmental driver of the COVID-19 outbreak in China. Lower and higher temperatures might be positive to decrease the COVID-19 incidence. M-SEIR models help to better evaluate environmental and social impacts on COVID-19.

**What is already known on this topic:**

- Many infectious diseases present an environmental pattern in their incidence.
- Environmental factors, such as climate and weather condition, could drive the space and time correlations of infectious diseases, including influenza.
- Severe acute respiratory syndrome coronavirus 2 (SARS-CoV-2) can be transmitted through aerosols, large droplets, or direct contact with secretions (or fomites) as influenza virus can.
- Little is known about environmental pattern in COVID-19 incidence.

**What this study adds:**

- The significant association between COVID-19 daily incidence and temperature was confirmed, using 3 methods, based on the data on COVID-19 and weather from 31 provincial-level regions in mainland China.
- Environmental factors were considered on the basis of SEIR model, and a modified susceptible-exposed-infectious-recovered (M-SEIR) model was developed.
- Simulations of the COVID-19 outbreak in Wuhan presented similar effects of temperature on incidence as the incidence decrease with the increase of temperature.

## INTRODUCTION

In December 2019, an outbreak of novel coronavirus pneumonia occurred in Wuhan, Hubei Province, China, and then were declared as an international public health emergency by the World Health Organization (WHO) on January 30 2020. The disease was officially named as coronavirus disease 2019 (COVID-19) and the newly emerged virus was named as SARS-CoV-2 in February 2020.^1^

Previous studies on early cases showed that the disease severity of COVID-19 with a 2.3% case-fatality rate,^2^ is much lower than Middle East Respiratory Syndrome (MERS) and Severe Acute Respiratory Syndrome (SARS).^3^ However, as Li et al. reported,^4^ the number of COVID-19 cases doubled every 7.4 days between December 2019 and January 2020, indicating COVID-19 might be more infectious than SARS and MERS. In March 2020, the outbreak of COVID-19 was declared as a global pandemic for the coronavirus rapidly expanded throughout China and to 116 other countries and territories worldwide.

Many infectious diseases present an environmental pattern in their incidence. A few studies on environmental issues, such as climate and weather condition, indicated that environmental factor could drive the space and time correlations of infectious diseases.^5-7^ Based on analysis on climate predicators, James D et al. found that humidity and temperature are optimal indicators in predicting influenza epidemics in tropical regions.^8^ Temperate regions of the Northern and Southern Hemispheres are characterized by highly synchronized annual influenza circulations during their winter months respectively.^5 7 8^ In the United States, an epidemiological study indicated that lower specific humidity is related to the occurrence of pandemic influenza, which is consistent with earlier finding in laboratory experiments.^9^ Absolute humidity, the actual mass of water vapor, is identified as a main cause of seasonal influenza epidemics.^10^ The influenza presents significant seasonal fluctuation in temperate monsoon climate regions as the absolute humidity varies greatly in summer and winter, which could help the multiplication of virus.

Severe acute respiratory syndrome coronavirus 2 (SARS-CoV-2) can be transmitted through aerosols, large droplets, or direct contact with secretions (or fomites) as influenza virus can.^11^ However, the environmental pattern remains to be elucidated in COVID-19 incidence. Based on dynamical equations, susceptible-exposed-infectious-recovered (SEIR) modeling has been developed and used to estimate key epidemic parameter to better characterize mechanism for the epidemic dynamics.^12-14^ Therefore, we explored the association between daily incidence and climate conditions using locally weighted regression and smoothing scatterplot (LOESS) and distributed lag nonlinear models (DLNMs) based on the data on COVID-19 and weather from 31 provincial-level regions in mainland China, between Jan 20 and Feb 29, 2020. Furthermore, we took account of environmental factors on the basis of SEIR model, and developed a modified susceptible-exposed-infectious-recovered (M-SEIR) model to characterize the climate impacts on epidemic dynamics.

## METHODS

### Study data

Data on COVID-19, including the number of new confirmed and probable cases were obtained from the China National Health Commission (CNHC) using the CoV2019 package^15^ (http://www.nhc.gov.cn/). COVID-19 data were collected among all of the 31 provincial-level regions in mainland China and Wuhan city, between Jan 20 and Feb 29, 2020.COVID-19 emerged in Wuhan city at the end of 2019 and rapidly spread across mainland China. Thus, population dynamic factors, including birth rate and death rate, were not considered here. Finally, daily incidences among the 31 provincial-level regions and Wuhan city were calculated by dividing the number of new confirmed cases by the population size at the end of 2018 respectively, and was reported per 100,000 population.

Daily temperatures (T) and relative humidity (RH) of 344 cities of the corresponding period were collected from the meteorological authority in mainland China. Means of temperatures and absolute humidity were further calculated for every provincial-level region. The Clausius-Clapeyron relation equation was used to calculate absolute humidity (AH) as following:

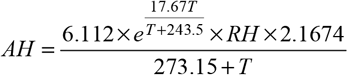

Data on climate conditions and population were retrieved from official reports previously released in mainland China. Therefore, the ethical review was not required.

### Statistical analysis

Trends of climate factors and daily COVID-19 incidence indicators, including the incidence and the common logarithm of numbers of newly confirmed cases (lgN), were analyzed with locally weighted regression and smoothing scatterplot (LOESS) in 31 provincial-level regions in mainland China from Jan 20 to Feb 29, 2020.

Developed on the definition of a cross-basis, DLNMs were used to infer the exposure-lag-response associations between climate factors and daily confirmed cases of COVID-19. DLNMs were constructed for mainland China outside of Hubei Province, Hubei Province outside of Wuhan city, and Wuhan city respectively. To induce the redundant analysis, temperature and absolute humidity of mainland China were represented by data on the capital, Beijing. Additionally, temperature and absolute humidity means of the sites in Hubei Province outside of Wuhan, were calculated as a representative of Hubei Province data.

To better understand the potential environmental driver of COVID-19, we took account of environmental factors on the basis of SEIR model and constructed the M-SEIR model to simulate the COVID-19 outbreak dynamic in Wuhan after travel restriction was put into force. Further sensitivity analysis was performed for quantitative risk assessment to evaluate the relationships between environmental parameter and COVID-19 incidence.

The equations of M-SEIR model were given in the following:

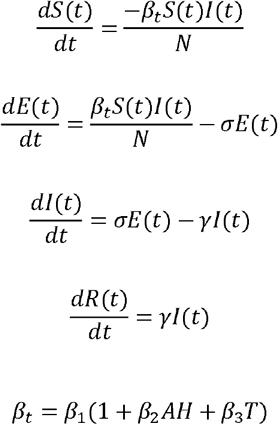

where S(t), E(t), I(t), and R(t) were the number of susceptible, exposed, infectious, and removed individuals at time t; 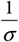 and 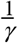 were the mean latent and infectious period; *β* was a time dependent rate of infectious contact; *β*_1_,*β*_2_ and *β*_3_ were constant coefficients.

The simulations of COVID-19 dynamic and sensitivity analysis were conducted by using the system dynamic section in AnyLogic software (version 8.5.2). The specific depict of parameter values in modified model and basic model details were included in **Supplementary Table 1**.

## RESULTS

80,981 cases of COVID-19 (cases of decrease in accounting not removed) was confirmed in 31 provincial-level regions in mainland China, between Jan 20 and Feb 29, 2020. Out of 80,981 cases, 68,034 (84.01%) were diagnosed in Hubei Province. Daily number of new confirmed cases and daily incidence in mainland China were presented in **Figure 1** and **Supplementary Table 1**. Daily number of cases peaked on Feb 12 and then it decreased, due to the adjustment in the diagnostic criteria of Hubei Province. And the number of cases and the incidence in China (outside of Hubei Province) have begun to decline early in Feb.

**Figure 1.**
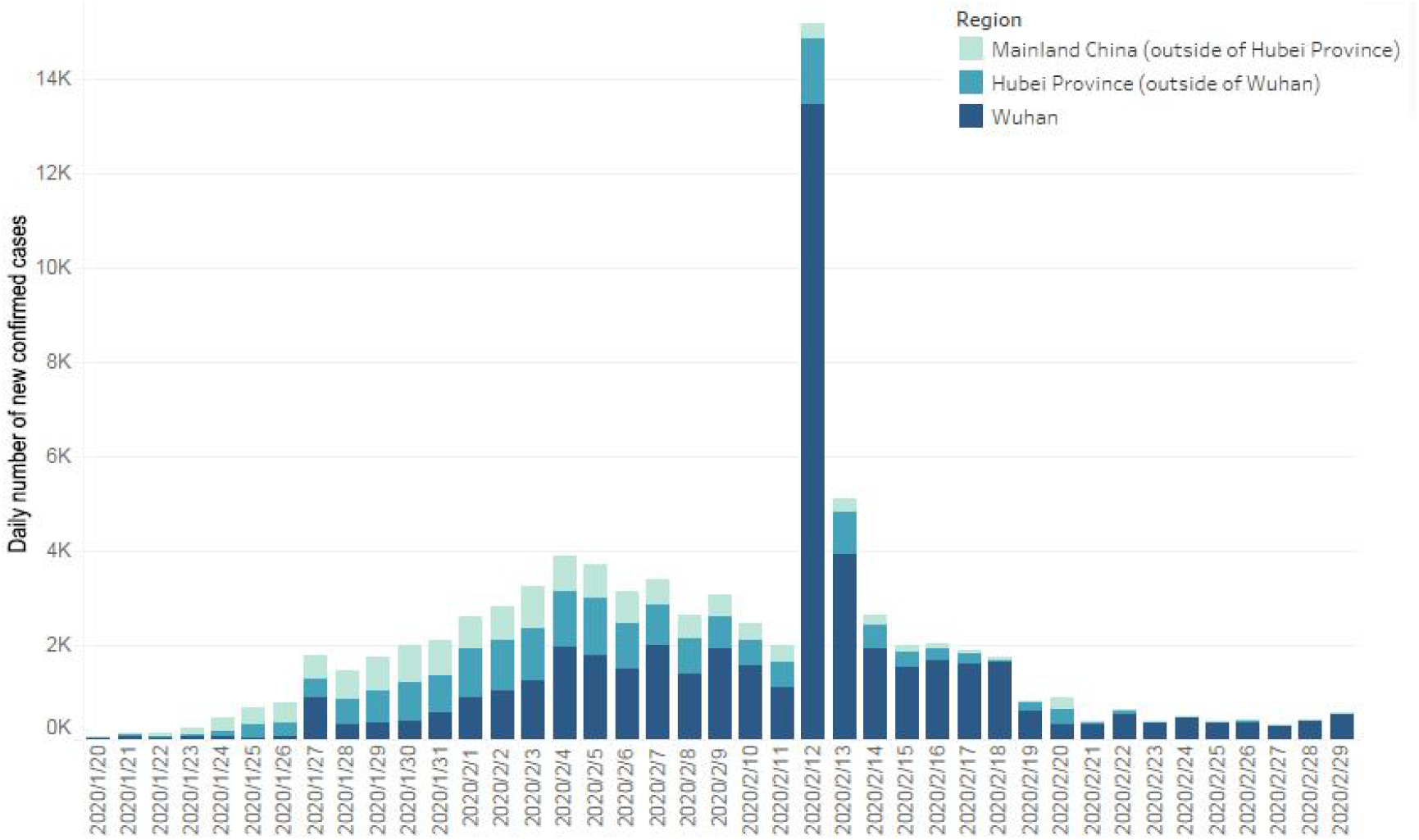
Daily number of new confirmed cases of COVID-19 in mainland China between Jan 20 and Feb 29, 2020.

From Jan 20 to Feb 29, 2020, temperature and absolute humidity varied in 31 provincial-level regions in mainland China (**Figure 2**). The highest temperature (26 °C) and absolute humidity (19.45 g/m^3^) were observed in Hainan Province and the lowest temperature (-22 °C) and absolute humidity (0.54 g/m^3^) were observed in Jilin Province, which resulted from the geographical location. COVID-19 daily incidence indicators (daily incidence and lgN) increased as the absolute humidity rose and declined slightly when absolute humidity reached approximately 7 g/m^3^ (**Figure 3**). Analysis for Hubei Province (outside of Wuhan) and Wuhan showed highly similar results (**Supplementary Figure 1 and Supplementary Figure 2**). Differences lay in the fact that cases clinically diagnosed without nucleic acid testing had been counted as confirmed cases in Hubei Province since Feb 12, which might increase potential bias in the model.

**Figure 2.**
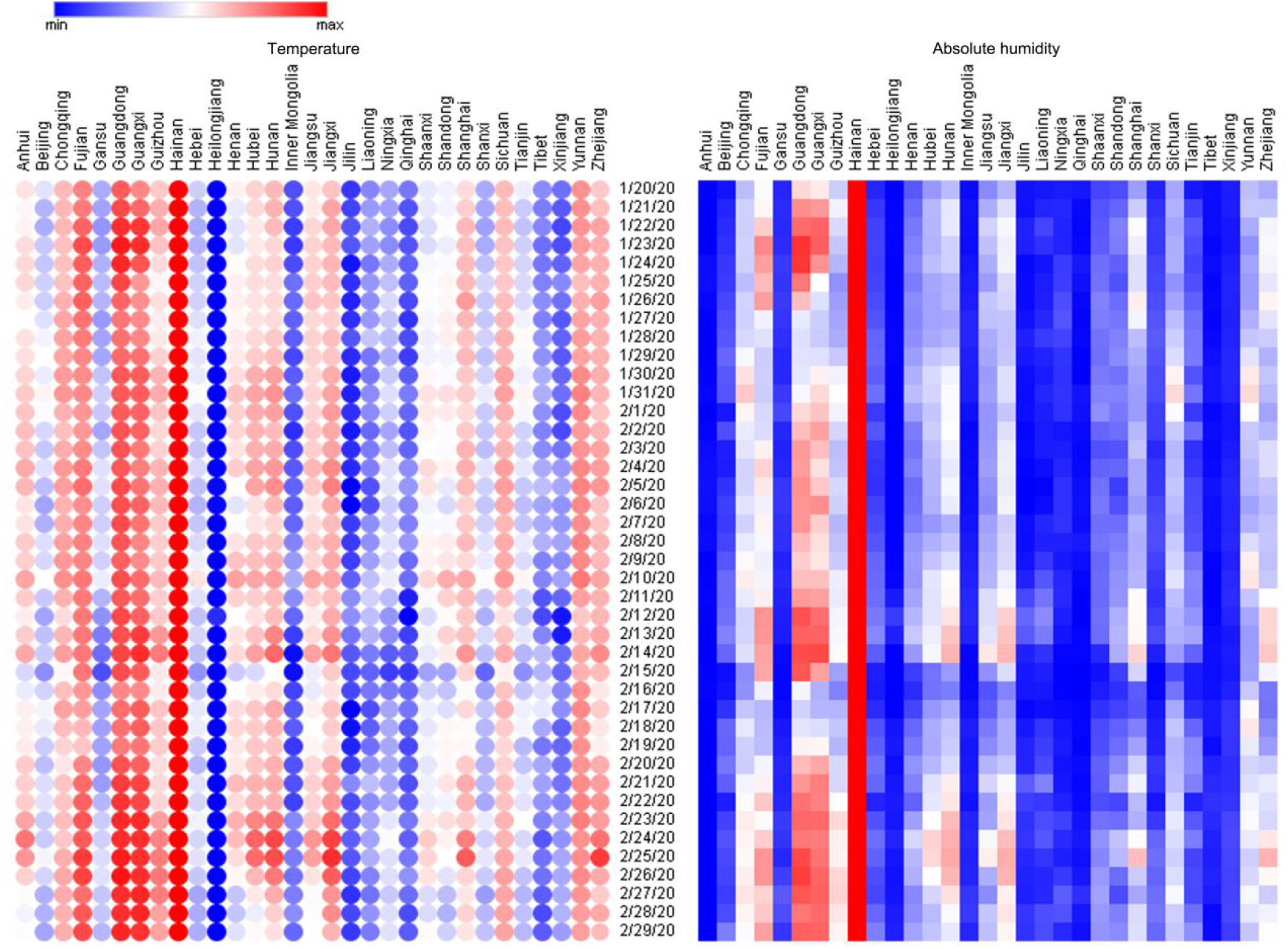
Between Jan 20 and Feb 29, 2020, temperature values (left columns) and absolute humidity values (right columns) in 31 provincial-level regions in mainland China.

**Figure 3.**
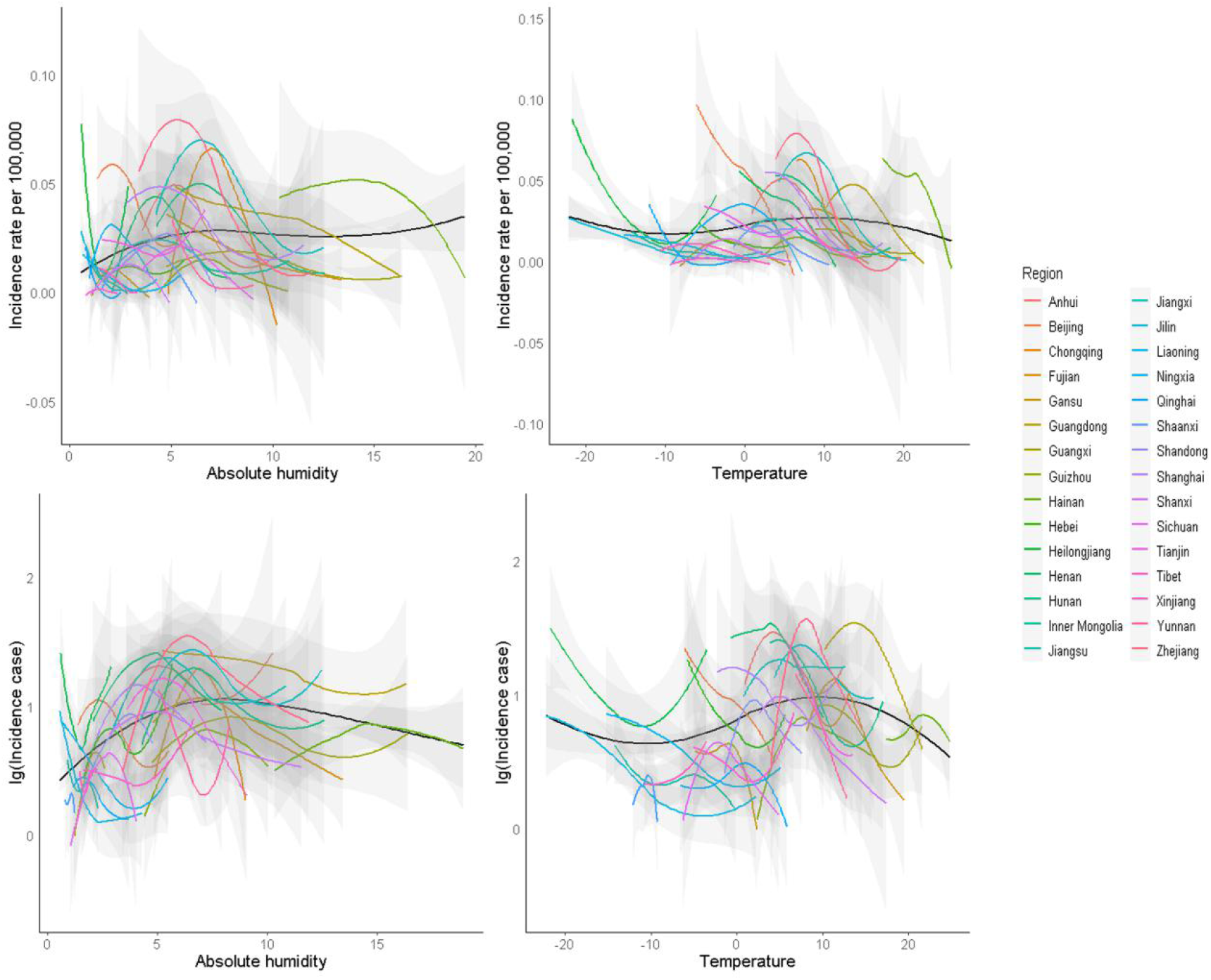
COVID-19 daily incidence indicators (daily incidence and lgN) and the expected values based on the temperature and absolute humidity in mainland China (outside of Hubei Province) from Jan 20 to Feb 29, respectively. The black line represents the expected value of a daily incidence and lgN based upon a LOESS regression for all days of available estimates. LOESS, locally weighted regression and smoothing scatterplots.

Associations between temperature and COVID-19 relative risk (RR) in mainland China (outside of Hubei Province), Hubei Province (outside of Wuhan) and Wuhan were presented as three-dimensional plots in **Figure 4**, compared with a reference value of 0 °C. The plots showed significant effect on COVID-19 incidence of temperature. In mainland China (outside of Hubei Province), the highest RR (1.71, 95% CI: 1.28-2.27) was observed at a cold temperature (-6 °C), suggesting the COVID-19 incidence were most likely to increase at -6 °C. The RR of 0.59 (95% CI: 0.44-0.78) at 6 °C rose to 1.06 (95% CI: 0.96-1.18) when temperature dropped to -6 °C. However, no statistical significance was found in lag-specific relative risk at lag 2 to lag 4, suggesting no delayed effect at any temperature. For example, the relative risk maintained at lag 2-4, as lag-specific RR was 1.14 (95% CI: 0.90-1.44) at lag 2 and 1.03 (95% CI: 0.86-1.33) at lag 4 when temperature was -6 °C. In Hubei Province (outside of Wuhan), RR was significantly higher at 8°C (RR 1.22, 95% CI: 1.07-1.38) and 10 °C (RR 1.92, 95% CI: 1.21-3.03) in lag 0. Conversely, lag-specific RR ranged from lag 0 to lag 7 at 8-10 °C, suggesting positive delayed effect on decreasing COVID-19 incidence during the condition. In Wuhan city, the highest RR 1.04 (95% CI: 0.92-1.17) without significance was observed at approximately 9 °C. However, the incidence was more likely to decrease with immediate and delayed effect at a lower or higher temperature than 9 °C. For example, RR was in a range of 0.64 (95% CI: 0.46-0.87) to 0.88 (95% CI: 0.73-0.99) at lag 0 to 5 days when the temperature was 4 °C and similar results were observed when the temperature was 16 °C.

**Figure 4.**
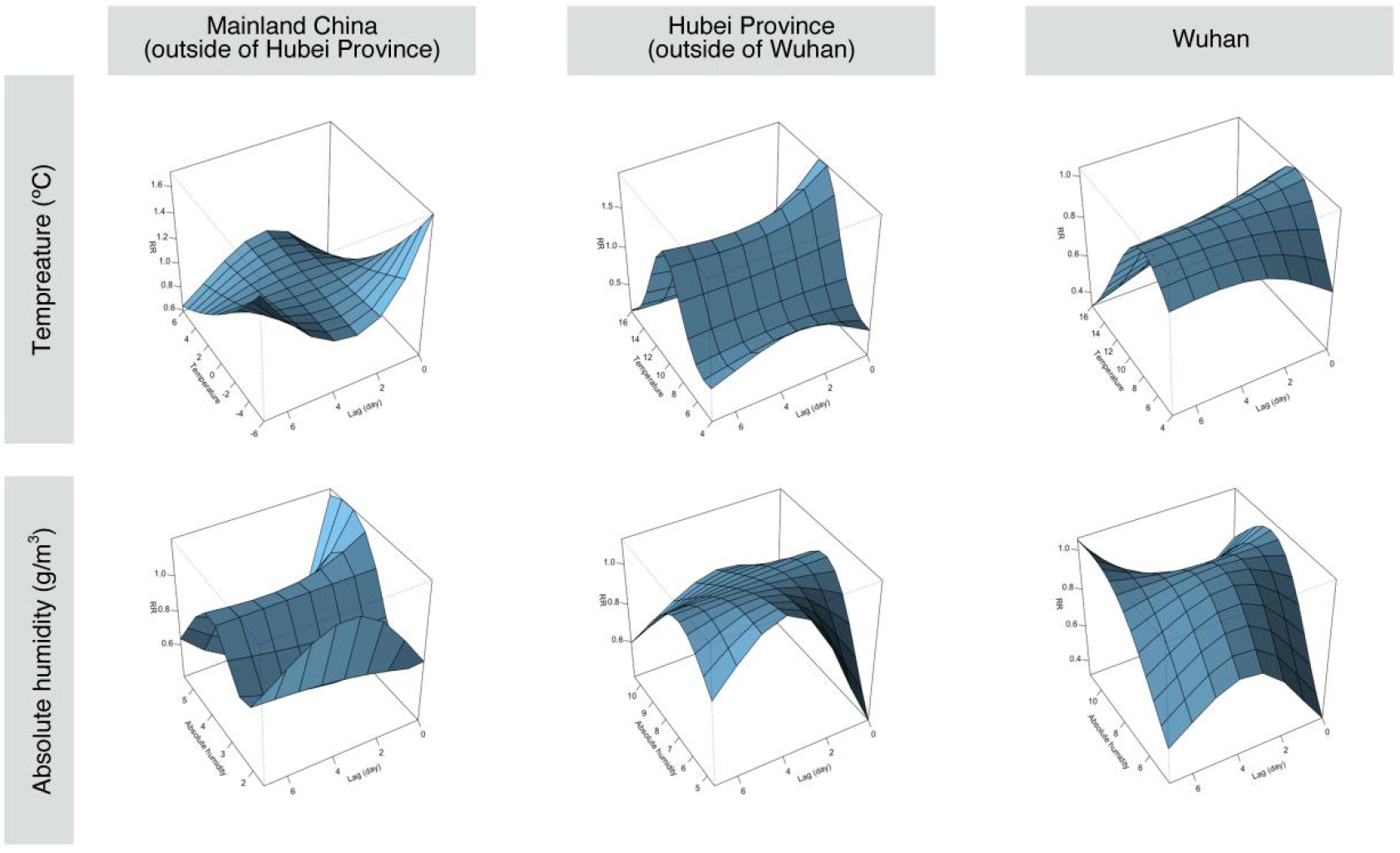
3-D plot of RR of COVID-19 along climate factors (temperature and absolute humidity) and lags in mainland China (outside of Hubei Province), Hubei Province (outside of Wuhan), and Wuhan city.

Overall pictures of the effect of absolute humidity on incidence in mainland China (outside of Hubei Province), Hubei Province (outside of Wuhan) and Wuhan were presented in **Figure 4**, showing 3-D graphs of COVID-19 relative risk (RR) along absolute humidity and lags compared with a reference value of 7.5 g/m^3^. The plots showed inconsistent effect of absolute humidity on COVID-19 incidence. In mainland China, immediate effect on COVID-19 incidence was strongest at absolute humidity of 4 g/m^3^ (RR: 1.13, 95%CI:1.02-1.27), indicating COVID-19 incidence was more likely to increase during the condition. When absolute humidity rose to 5 g/m^3^, values of lag-specific RR were in range of 0.60 (95% CI: 0.36-0.99) to 0.62 (95% CI: 0.41-0.93) at lag 3 to lag 5 (**Supplementary Figure 3**), indicating a strong delayed effect on COVID-19 incidence at absolute humidity of 5 g/m^3^. In Hubei Province, immediate effect on reducing COVID-19 incidence was observed when absolute humidity ranged from 4.5 g/m^3^ (RR 0.40, 95% CI: 0.19-0.84) to 5.5g/m^3^ (RR 0.65, 95% CI: 0.44-0.96) (**Supplementary Figure 4**). However, no significant difference was observed in absolute humidity in Wuhan city (**Supplementary Figure 5**).

**Figure 5.**
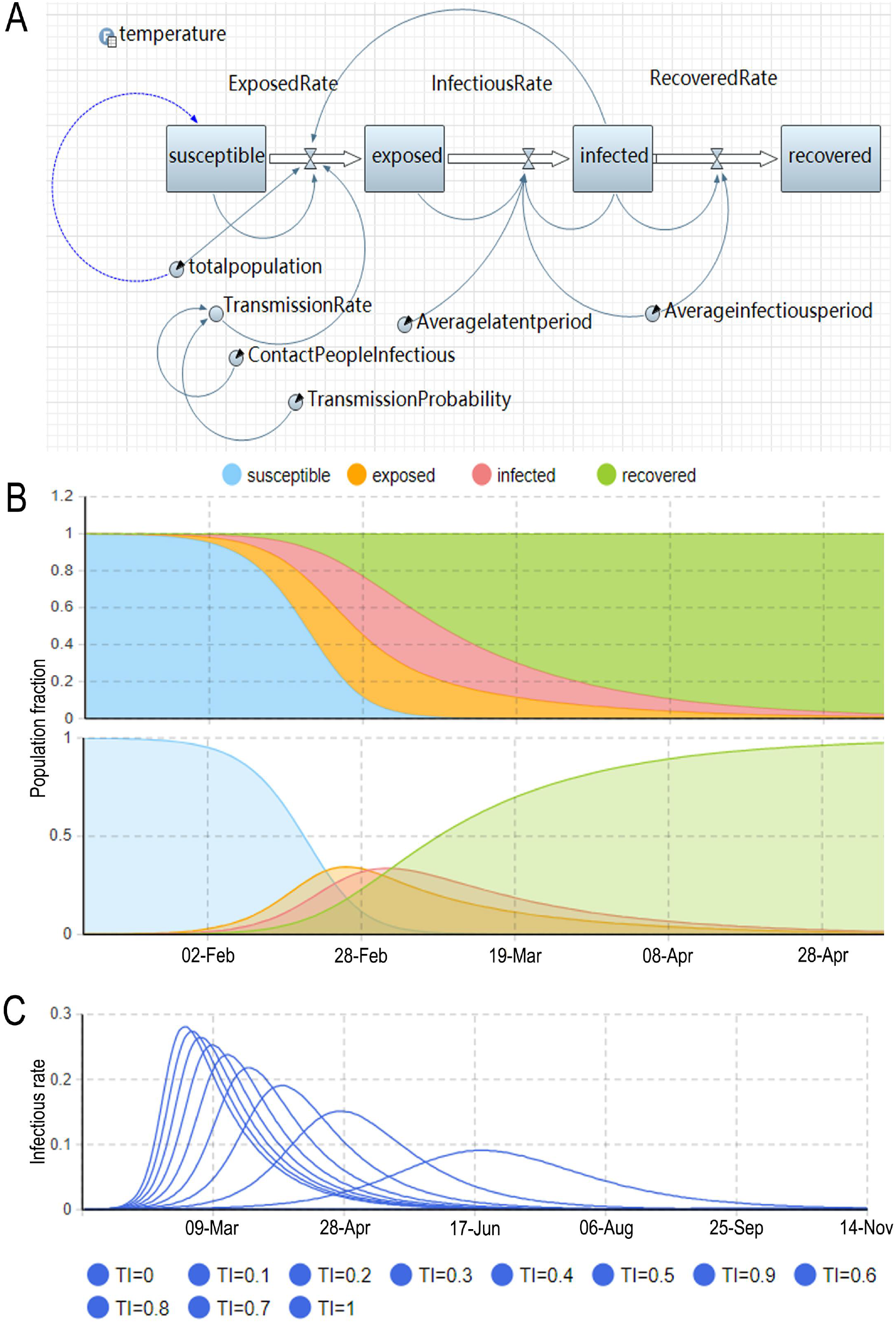
COVID-19 dynamic trends and sensitivity analysis using M-SEIR model in Wuhan. **(A)** The over-all structure of M-SEIR model constructed by using the system dynamic section in AnyLogic software. **(B)** The snapshot represents the different population proportion of susceptible, exposed, infected, and recovered states under the specific time-point and forecasts the trend of the COVID-19 outbreak in Wuhan city. **(C)** Sensitivity analysis under different temperature scenarios in Wuhan city. As the temperature-corrected transmission index rises, the peak of the curve increased under different times gradually. M-SEIR model, modified susceptible-exposed-infectious-recovered model; TI, the temperature-corrected transmission index (i.e. The transmission rate for susceptible to exposed, *β*_t_).

Considering the environmental impacts, we constructed the M-SEIR model to simulate the dynamic of COVID-19 by using the system dynamic section in AnyLogic software. SEIR dynamic transmission model compartmentalized the population into four states including susceptible, exposed, infected, and recovered, and further analyzed the relationships and interconnection using stock and set parameters, flows and table function (**Figure 5A; Supplemental video 1**). We set the initial values of the parameter and incorporated the temperature index in Wuhan city from Jan 20 and Feb 29, 2020, into the modified SEIR model. **Supplemental table 3** presented the comparison of modified SEIR model in our study and classic SEIR models in similar studies. The four curves were stratified by types of state, and showed a similar pattern: the population size increased early in epidemic and then decreased as the period ends (e.g., due to recovery). As the M-SEIR model predicted, the number of infections would peak around Mar 5, reaching the inflection point, and the COVID-19 outbreak in Wuhan would be expected to end by late April (**Figure 5B; Supplemental video 1**). Furthermore, a sensitivity analysis on the transmission rate adjusted by temperature indicated high stability of our

M-SEIR model (**Figure 5C; Supplemental video 2**). We set the transmission rate from 0 to 1 with a step of to 0.1, and conducted the simulations to reduce the bias involving in the model, parameters, and functional relationships. Finally, we found that the transmission rate decreased with the increase of temperature, leading to the decrease of infection rate and outbreak size.

## DISCUSSION

We inferred that the number of new confirm COVID-19 cases in mainland China peaked on Feb 1, 2020. COVID-19 daily incidence were lowest at -10 °C and highest at 10 °C,while the maximum incidence was observed at the absolute humidity of approximately 7 g/m^3^. We found significant association between temperature and COVID-19 daily incidence due to the immediate and delayed effect observed using DLNMs. As predicted in M-SEIR model, the COVID-19 outbreak would peak around March 5, 2020 and end in late April in Wuhan. Additionally, we found that transmission rate decreased with the increase of temperature, leading to further decrease of infection rate and outbreak size. Therefore, temperature drive the space and time correlations of COVID-19, and it can be used as an optimal predicator.

In this study, we inferred the significant association between temperature and COVID-19 daily incidence using LOESS, DLNMs and M-SEIR model, suggesting that temperature plays an important role in the outbreak of COVID-19 and can be used in predicting the potential spread of COVID-19. Lower and higher temperatures may be positive to decrease the COVID-19 incidence, which help to shed new light on the environmental drivers of COVID-19 in China. Our results are in line with the findings in SARS. Based on data on SARS and climate in 4 cities, Tan et al. found that temperature is a powerful indicator for SARS-CoV transmission, in which the risk of increased daily incidence differed between the effects of high and low temperatures.^16^ Additionally, Lowen’s laboratory work evidenced that temperature affect the virus spread of aerosol using a guinea pig model.^17^ However, the temperature DLNM in Hubei Province, showed different patterns from those in mainland China and Wuhan city, as COVID-19 relative risk rose at a moderate temperature.

In our analysis, we failed to observe a significant relationship between absolute humidity and COVID-19 incidence based on the data of mainland China. However, absolute humidity has been reported as a strong correlation with influenza epidemic, due to the seasonal pattern that influences the multiplications and spread of influenza.^9 18 19^ In another study on MERS, caused a lethality of more than 35%, confirmed that the activity of MERS-CoV in droplet or aerosol, decreases significantly as absolute humidity increases though the mechanism is not yet clear.^20^ The difference between our study and previous finding may result from the fact that absolute humidity remained stable in a region during a very limited period. Additionally, rapid and strong actions taken by the government could biased our study. Despite of the negative consequence in our study, further studies on absolute humidity are required to perform.

Combination of infectious disease dynamics model and environmental patterns is required to better explain the relationship between environmental factors and infection.^21^ Dynamic transmission model was usually performed to predict the genesis and development trend of infectious diseases as well as to evaluate the effect of intervention but few dynamic transmission models included environmental factors for the increasing uncertainty. However, to reveal the dynamic of an infectious disease, it would be much better to take account of environmental impact on the basis of dynamic transmission model.^22 23^

Environmental factors, characterized by lag effects and threshold effects, can target at two objects, host and virus, during infectious disease outbreak. On one hand, human activity patterns and immunity can be influenced by environmental factors. But the effect caused by environmental condition was limited during the COVID-19 outbreak, due to the absence of extreme weather and specific immunity for a newly emerging virus. On the other hand, environmental impacts on the SARS-CoV-2 are more significant than the host population because the transmission and virulence of the virus varies in different conditions. Finally, environmental impacts on transmission of virus should be characterized in the dynamic model, because infectiousness estimated in the traditional dynamic model is actually a confounding effect with environmental effect. It is necessary to take account of environmental issues on the basis of dynamic transmission model so that the impacts could be isolated and qualified. A dynamic model is not only compatible with the infectious disease transmission mode for virus itself, but also can be well coupled with surveillance data on environmental issues.^24^ Consequently, we constructed a M-SEIR model to correct the potential deviation of temperature to simulate the dynamic epidemic of COVID-19. The M-SEIR model predicted that the outbreak would reach its peak reach an inflection point around March 5, 2020, which is consistent with the actual situation based on data released by the NHC.^25-29^ And it is expected that the COVID-19 outbreak in Wuhan would end in late April. In addition, we conducted a sensitivity analysis on the temperature-adjusted transmission rate. Finally, we found transmission rate decreased with the increase of temperature, leading to further decrease of infection rate and epidemic size.

Our analysis is subject to limitations. First, the COVID-19 dynamics are determined by multiple factors, including virus, climate, socio-economic development, population mobility, population immunity, and urbanization. However, not all those factors were considered in this study. Second, the parameters of M-SEIR models were optimized, based on the previous analysis which might be biased by the lack of official data and the adjustment of diagnostic criteria in the outbreak. Third, it’s an ecological analysis in very short period so that we cannot avoid the bias caused by other ecological factors changed over time.

### Conclusions and public health implications

Temperature is an environmental driver of the COVID-19 outbreak in China. Lower and higher temperatures might be positive to decrease the COVID-19 incidence. As predicted in M-SEIR model, the COVID-19 outbreak would peak around March 5, 2020 and end in late April in Wuhan. Modified-SEIR models help to better evaluate and identify national and international prevention and intervention targeted COVID-19. The COVID-19 outbreak would not last for a long period of time with the increase of temperature, but the scale of the outbreak would be influenced by the measures taken among countries.

## Contributions

All authors contributed to the study concept and design. PS, HY, and XL wrote the paper. PS and YD collected the data. PS, YD and CZ analyzed the data. PS, HY, ST, WL, MH, and SX reviewed and revised the manuscript before submission. All authors approved the final submitted version.

## Funding

The work was supported by the National Key Research and Development Program of China to Prof Shuhua Xi (2018YFC1801204). The funders had no role in design and conduct of the study; collection, management, analysis, and interpretation of the data; preparation, review, and approval of the manuscript; or the decision to submit the manuscript for publication.

## Competing interests

All authors have completed the ICMJE uniform disclosure form at http://www.icmje.org/coi_disclosure.pdf (available on request from the corresponding author) and declare: no support from any organization for the submitted work other than those described above; no financial relationships with any organizations that might have an interest in the submitted work in the previous three years; no other relationships or activities that could appear to have influenced the submitted work.

## Transparency declaration

The manuscript’s guarantor affirms that the manuscript is an honest, accurate, and transparent account of the study being reported; that no important aspects of the study have been omitted; and that any discrepancies from the study have been explained.

## Data sharing

No additional data available.

## Notes

### Competing Interest Statement

The authors have declared no competing interest.

